# Power law behaviour in the saturation regime of fatality curves of the COVID-19 pandemic

**DOI:** 10.1101/2020.07.12.20152140

**Authors:** Giovani L. Vasconcelos, Antônio M. S. Macêdo, Gerson C. Duarte-Filho, Arthur A. Brum, Raydonal Ospina, Francisco A. G. Almeida

## Abstract

We apply a versatile growth model, whose growth rate is given by a generalised beta distribution, to describe the complex behaviour of the fatality curves of the COVID-19 disease for several countries in Europe and North America. We show that the COVID-19 epidemic curves not only may present a subexponential early growth but can also exhibit a similar subexponential (power-law) behaviour in the saturation regime. We argue that the power-law exponent of the latter regime, which measures how quickly the curve approaches the plateau, is directly related to control measures, in the sense that the less strict the control, the smaller the exponent and hence the slower the diseases progresses to its end. The power-law saturation uncovered here is an important result, because it signals to policymakers and health authorities that it is important to keep control measures for as long as possible, so as to avoid a slow, power-law ending of the disease. The slower the approach to the plateau, the longer the virus lingers on in the population, and the greater not only the final death toll but also the risk of a resurgence of infections.

## 1 Introduction

The COVID-19 pandemic has been recognised as one of the gravest public health crises in recent history. At the time of writing, more than 15 million infections have been confirmed worldwide and over 620,000 deaths have been attributed to the disease. Although many countries, notably in Far-East Asia and Europe, have been able to control somewhat the spread of the disease, the SARS-CoV-2 virus is still advancing in many parts of the world—an alarming situation that prompted the WHO Director-General to state on July 9, 2020, that “in most of the world the virus is not under control. It is getting worse.”^1^ Furthermore, countries that have gained some control over the epidemic are not entirely safe, as there persists a potential risk of resurgence of infections, since a large percentage of their population remains susceptible to the virus.

It is therefore important for governmental and health authorities not only to adopt measures to contain the early spread of the virus, but also to keep those measures in place, in one way or another, for as long as possible, even after the worst phase of the epidemic has been overcome, so as to bring the advance of the disease to a near halt as quickly as possible. In this context, where the epidemic remains a danger from beginning to end, it is important to have models that can give a full description of the epidemic dynamics: from its early rapid-growth phase to the late saturation regime, in which the cumulative number of cases or deaths tends to a levelling plateau.

Epidemics, in general, and the COVID-19 pandemic, in particular, exhibit a complex dynamics that results from the intertwined interaction between the virus’s modus operandi and the human responses: the virus propagates in a complex network of human contacts, which is affected by containment measures (or lack thereof), which in turn impacts back on the virus rate of spread, and so on. Power-law behaviour is a distinct signature of complexity, and in such a guise they are expected to be present in epidemic dynamics as well. Evidences of complexity in the COVID-19 epidemic dynamics have indeed been observed in a number of recent works, most noticeably in the form of a power-law behaviour in the early growth regime of case and death curves^2–6^, but also in the long-time asymptotic of the probability to become infected^7–9^. It has also been pointed out that such complex behaviour lies outside the range of applicability of standard SIR-type models, and a number of alternative dynamics have been suggested, such as the recent proposal of a random walk on a hierarchic landscape of social clusters^10–12^.

Phenomenological growth models offer a simplified but powerful tool^13^ to describe the complexity of COVID-19 epidemic data. On the one hand, their simplicity stems from the fact they are formulated in terms of a single differential equation that often admits an exact analytical solution, which renders such models quite amenable to both mathematical analysis and numerical application to empirical data. On the other hand, their power derives from the fact that the model parameters seek to capture (in an effective manner) the basic aspects of the underlying epidemic dynamics, but without having to describe epidemiological mechanisms that may be difficult to identify^14^. Nevertheless, variations in the mechanisms of the disease spread, say, owing to demographics or economic factors as well as to different response interventions, will be naturally reflected in changes of the model parameters from group to group. Thus, when properly applied and interpreted, growth models can provide useful insights into the spread of novel infectious diseases^15^.

In this paper we apply a generalised logistic model^16^, which we refer to as the *beta logistic model* (BLM), to study the fatality curves of COVID-19, as represented by the cumulative number of deaths as a function of time, for different countries. As will be argued later, the BLM is one of the simplest growth models that is capable of capturing the three main generic features of a cumulative epidemic curve: i) an initial subexponential growth; ii) an intermediate linear regime; and iii) a final subexponential saturation trend toward the plateau; see Fig. 1 for a schematic of a generic epidemic curve. (Exponential behaviour can be attained in both early and late regimes as limiting cases.) We show that the BLM describes very well the mortality curves of the eight countries selected here to exemplify the model, namely: Belgium, Canada, France, Germany, Italy, Netherlands, United Kingdom, and United States. In particular we show that these countries exhibit a polynomially slow approach to the plateau, in the sense that the growth rate asymptotically decays in time as a power-law, rather than exponentially fast as predicted by mechanistic models of the SIR type.

**Figure 1.**
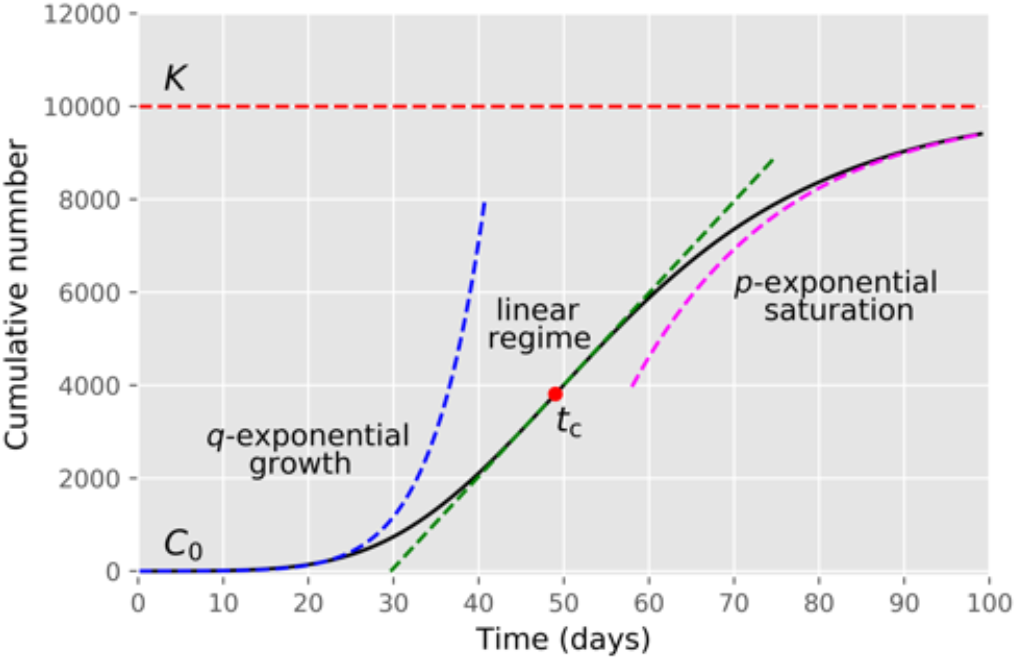
Schematic of a typical epidemic curve for the cumulative number of cases or deaths, where the three main regimes of growth are indicated: i) early *q*-exponential growth; ii) intermediate region of linear growth; and iii) *p*-exponential saturation towards the plateau. Here *C*_0_ is the value at *t* = 0, *K* is the value at the end of the epidemic (plateau), and *t*_*c*_ denotes the inflection point of the curve.

In this context, it is important to bear in mind that there is an intrinsic connection between mechanistic epidemic models of the SIR type and growth models. For instance, it is sometimes possible to map the entire curve of some growth models onto the corresponding curve of a SIRD-type model^17^. We remark, however, that given the polynomial nature of the early and late time evolution that we find in the analysed data, the target SIRD-type model, to be put in correspondence with the BLM, would have to be modified, for example, by the inclusion of a power law in the incidence term, as proposed by^18^, or by considering time-dependent parameters, as studied in^19^. This interesting possibility of connection between generalised SIRD models and the BLM is, however, beyond the scope of the present paper and is left for future studies.

The results reported in the present paper are important for several reasons. First, we show that countries that have kept stricter measures for longer periods tend to experience a faster approach to the plateau; whereas countries with less strict measures throughout the epidemic or that relaxed those measures at an earlier stage usually display a slow saturation regime. This slow progress to the end of the epidemic represents an extra danger, for the longer the virus lingers on in a population, the greater the risk of a resurgence of cases. Second, our results are also relevant from a more fundamental viewpoint, since they reveal a seemingly new power-law dynamical regime in the final stages of the COVID-19 epidemics. Third, we hope that our findings will stimulate further research on the problem of better characterising the final phase of epidemic dynamics.

## 2 Results

### 2.1 Mathematical Results

We model the time evolution of the number of cases (infections, deaths, etc) in the epidemic by means of the beta logistic model (BLM), defined by the following differential equation^16^:

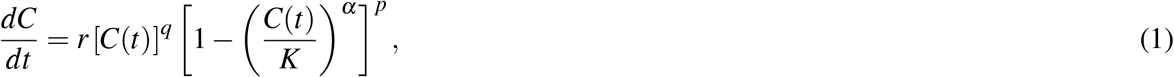

where *C*(*t*) is the cumulative number of cases at time *t, r* is the growth rate at the early stage, and *K* is the final epidemic size. The exponent *q* controls the initial growth regime and allows to interpolate from linear growth (*q* = 0) to subexponential growth (*q <* 1) to purely exponential growth (*q* = 1); whilst the exponent *p* controls the rate of approach to the plateau, with *p >* 1 implying a polynomial rate, whereas *p* = 1 yields an exponentially fast approach (see below). The exponent *α* is the so-called asymmetry parameter that controls the degree of asymmetry with respect to the symmetric S-shape of the logistic curve, which is recovered for *q* = *p* = *α* = 1, so that the larger the exponent *α*, the quicker the curve bends towards the plateau.

A detailed mathematical analysis of the BLM is left for the section Methods. Here it suffices to quote some important properties of the model. First, equation (1) can be integrated exactly to yield the following analytical solution in implicit form, in terms of the curve *t*(*C*):

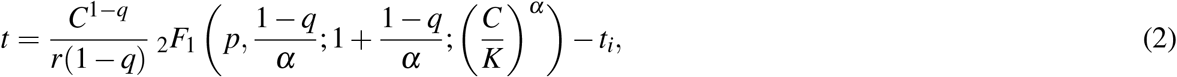

where _2_*F*_1_(*a, b*; *c*; *x*) is the Gauss hypergeometric function^20^ and

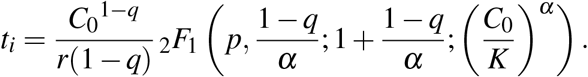

The fact that the solution of the BLM comes in implicit form does not represent any numerical impediment, as the above solution can easily be applied, say, for curve-fitting purposes by viewing the empirical data in the same ‘implicit’ form *t*(*C*). To the best of our knowledge, the exact solution for the BLM given in (2) has not appeared before in the literature. Exact solutions for certain particular cases have been obtained before^16,21^, but it appears that an analytical solution for arbitrary values of *q, p*, and *α*, as written in (2), was not known previously.

The inflection point of the curve *C*(*t*), which is defined as the point where the second derivative vanishes, i.e. 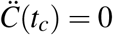, where dots denote time derivative, is given by

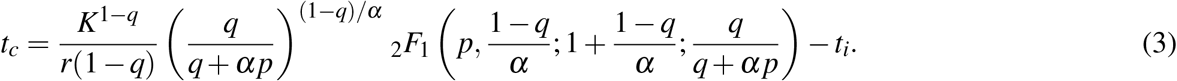

The small- and large-times asymptotic behaviour of *C*(*t*) are found to be as follows:

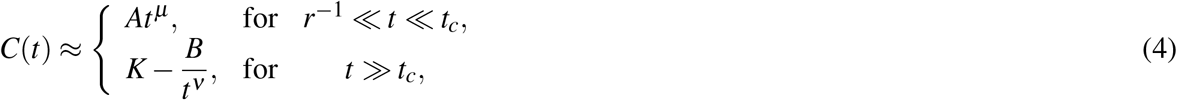

where *µ* = 1*/*(1*− q*), *A* = [*r*(1*− q*)]^1*/*(1*−q*)^, ν = 1*/*(*p−* 1), and *B* = [*K*^*p−q*^*/*(*p−*1)*rα*^*p*^]^1*/*(*p−*1)^. (For *q→*1 and *p→*1, one obtains exponential growth and exponential decay, respectively; see Subsection Mathematical Model.)

Recall that the inflection point *t*_*c*_ of the cumulative curve *C*(*t*) corresponds to the “peak” of the daily curve *Ċ*(*t*). From (4) we thus see that the daily number of cases predicted by the BLM has an early *polynomial growth* (i.e., before the peak) and a *power-law decay* in the saturation phase (i.e., after the peak):

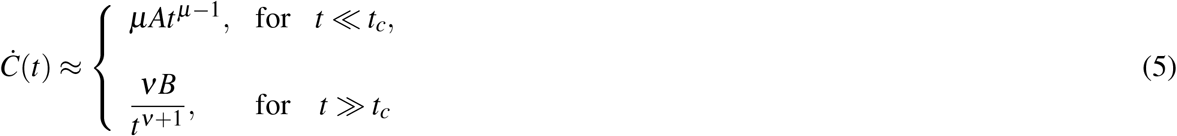

for *q <* 1 and *p >* 1.

The behaviour of *C*(*t*) near the inflection point *t*_*c*_ is given by

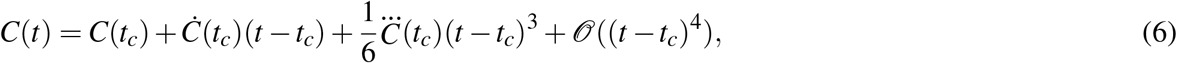

Where

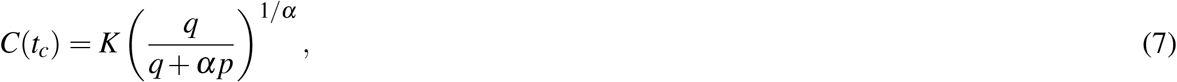

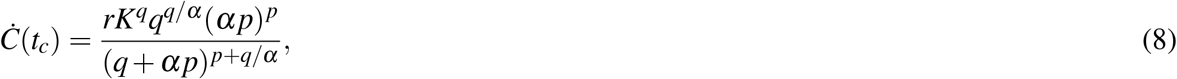

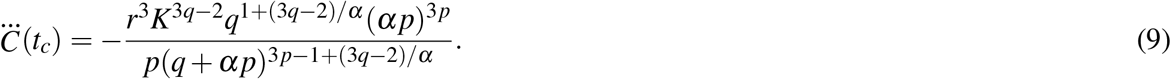

In particular note that *C*(*t*_*c*_) does not depend on the growth rate *r*, as expected, since *r* only sets the time scale of the problem. Furthermore, for large values of *α* and *p*, the growth ‘velocity’ *Ċ*(*t*_*c*_) at the inflection point becomes simply *Ċ*(*t*_*c*_) *≈ rK*^*q*^, which indicates that the slope of the curve *C*(*t*) at the inflection point is mainly controlled by *r*, together with *q* and *K*.,..but less dependent on both *α* and *p*. Similarly, for large *α*, the jerk (or jolt) 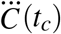 does not depend on *p*: 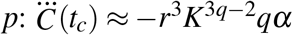, which confirms the fact anticipated above that (for fixed *r, q*, and *K*) the exponent *α* is the main responsible for controlling the ‘bending’ of the curve towards the plateau.

The above summarized analysis thus shows that each of the main features of an epidemic curve can be said to be mainly governed by a specific parameter of the BLM, as follows: i) *K* is, of course, the plateau height; ii) *q* determines the nature of the early subexponential growth; iii) *r* sets the time scale of the problem and hence it is the main responsible for determining the slope of the linear region; iv) *α* controls the sharpness of the bending towards the plateau; and v) *p* characterizes the power-law saturation regime.

### 2.2 Application to Empirical Data

As discussed above, the BLM is capable of capturing both a subexponential growth and a subexponential approach to the plateau (with exponential behaviors contemplated in the limits *q; p→*1). This thus makes the BLM suitable for situations where the epidemic has reached the intermediate to final stage, such that the corresponding cumulative epidemic curve has a well defined inflection point and is clearly trending toward a possible plateau. With this in mind, we have chosen countries whose cumulative curves of deaths due to COVID-19 seemed to be approaching a plateau up to the date of the present analysis. As representative examples of these situations, we have chosen the following countries for the present study: Belgium, Canada, France, Germany, Italy, Netherlands, United Kingdom, and United States. It is worth noting that several of these countries seem to have experienced a resurgence of infections in the late Summer of 2020. To avoid eventual ‘second wave’ effects, we have therefore restricted our data analysis up to July 31, 2020, except for the United States, where we used data up to July 7, 2020, because there a re-acceleration of the epidemic curve appeared earlier than in the other countries.

In the left panels of Figs. 2-5 we show the cumulative death curves (red circles) for the selected countries, together with the corresponding best fits (black solid curves) by the BLM. From these figures, one sees that the theoretical curves describe remarkably well the empirical data for all cases considered. The corresponding fitting parameters, together with their respective 95% Confidence Intervals (CI), are given in table 1.

**Table 1.** Parameter estimates of the BLM for the selected countries. with the respective 95% Confidence Intervals given in parenthesis.

| Country | $p$ | $q$ | $r$ | $\alpha$ | $K$ |
| --- | --- | --- | --- | --- | --- |
| Canada | 1.08 (1.05, 1.11) | 1.00 | 0.29 (0.27, 0.31) | 0.23 (0.20, 0.32) | 9081 (90709, 9104) |
| Italy | 1.09 (1.08, 1.09) | 1.00 | 1.24 (1.17, 1.68) | 0.06 (0.06, 0.14) | 35311 (35307, 35328) |
| United Kingdom | 1.14 (1.12, 1.15) | 1.00 | 0.76 (0.62, 0.89) | 0.11 (0.10, 0.16) | 41680 (41637, 41731) |
| Netherlands | 1.24 (1.18, 1.33) | 0.83 (0.73, 1.00) | 0.54 (0.38, 0.78) | 0.92 (0.28, 1.53) | 6198 (6192, 6206) |
| Germany | 1.34 (1.32, 1.37) | 1.00 | 0.36 (0.34, 0.37) | 0.35 (0.32, 0.48) | 9285 (9274, 9298) |
| Belgium | 1.38 (1.32, 1.46) | 0.91 (0.92, 1.00) | 0.38 (0.28, 0.50) | 0.85 (0.49, 1.24) | 9909 (9897, 9924) |
| France | 1.87 (1.69, 2.11) | 1.00 | 0.18 (0.17, 0.18) | 1.19 (1.01, 1.42) | 31151 (30923, 31472) |
| United States | 3.43 (2.66, 5.02) | 0.96 (0.96, 0.99) | 0.35 (0.33, 0.37) | 0.64 (0.50, 0.78) | 229249 (200507, 286172) |

**Figure 2.**
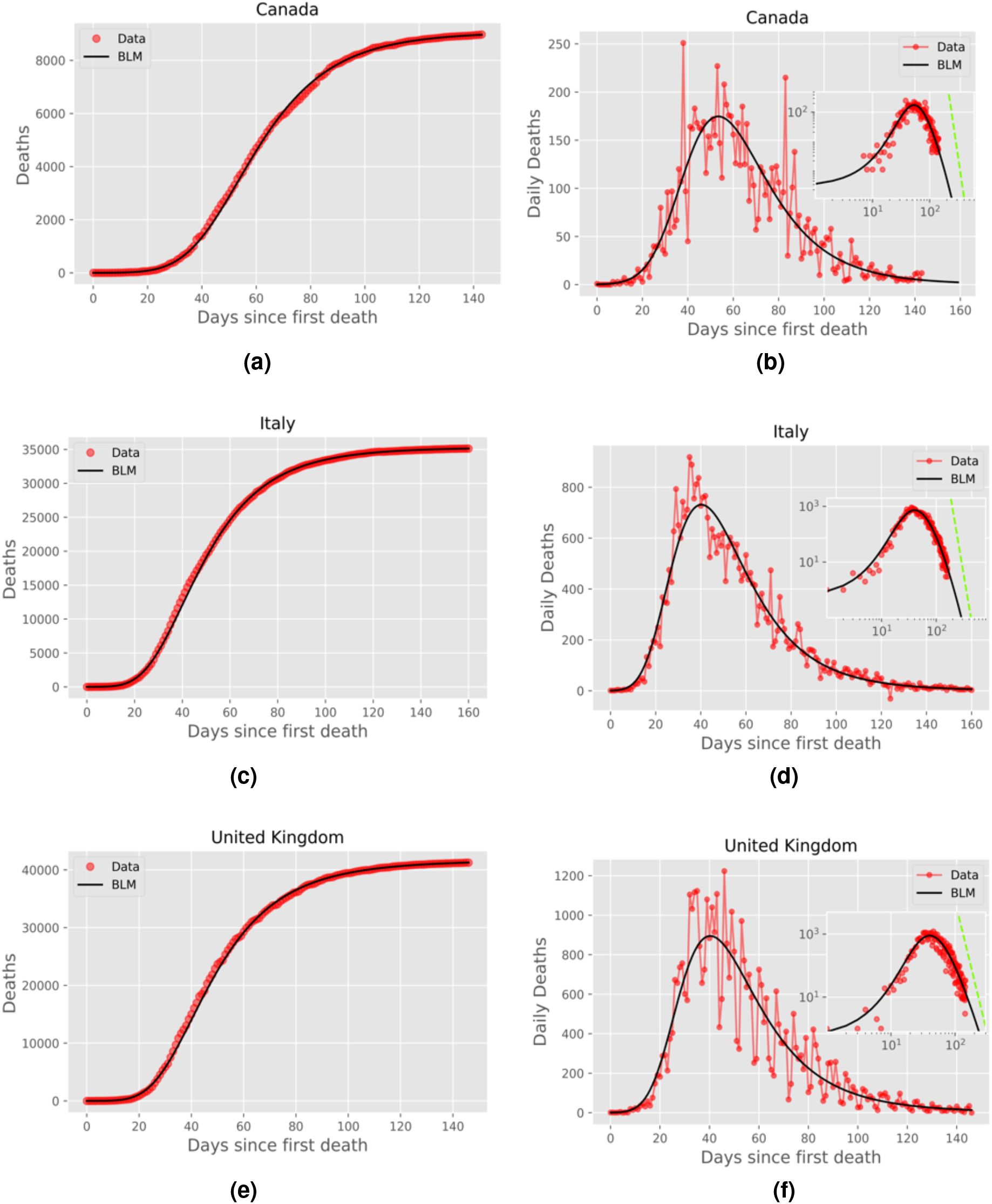
Left panels: Cumulative number of deaths (red circles) attributed to COVID-19 up to July 31, 2020, for (a) Canada, (c) Italy, and (e) United Kingdom; the solid black curves are the best fits by the beta logistic model, with the corresponding parameters given in table 1. Right panels: Daily number of deaths for the same countries, where the empirical data are indicated by red circles and the solid black curve represent the time derivative of the respective theoretical curve in the left panels; the insets show the same graphs as in the main plots, only in log-log scale, where the green straight lines correspond to the power-law asymptotic behavior predicted by (5).

**Figure 3.**
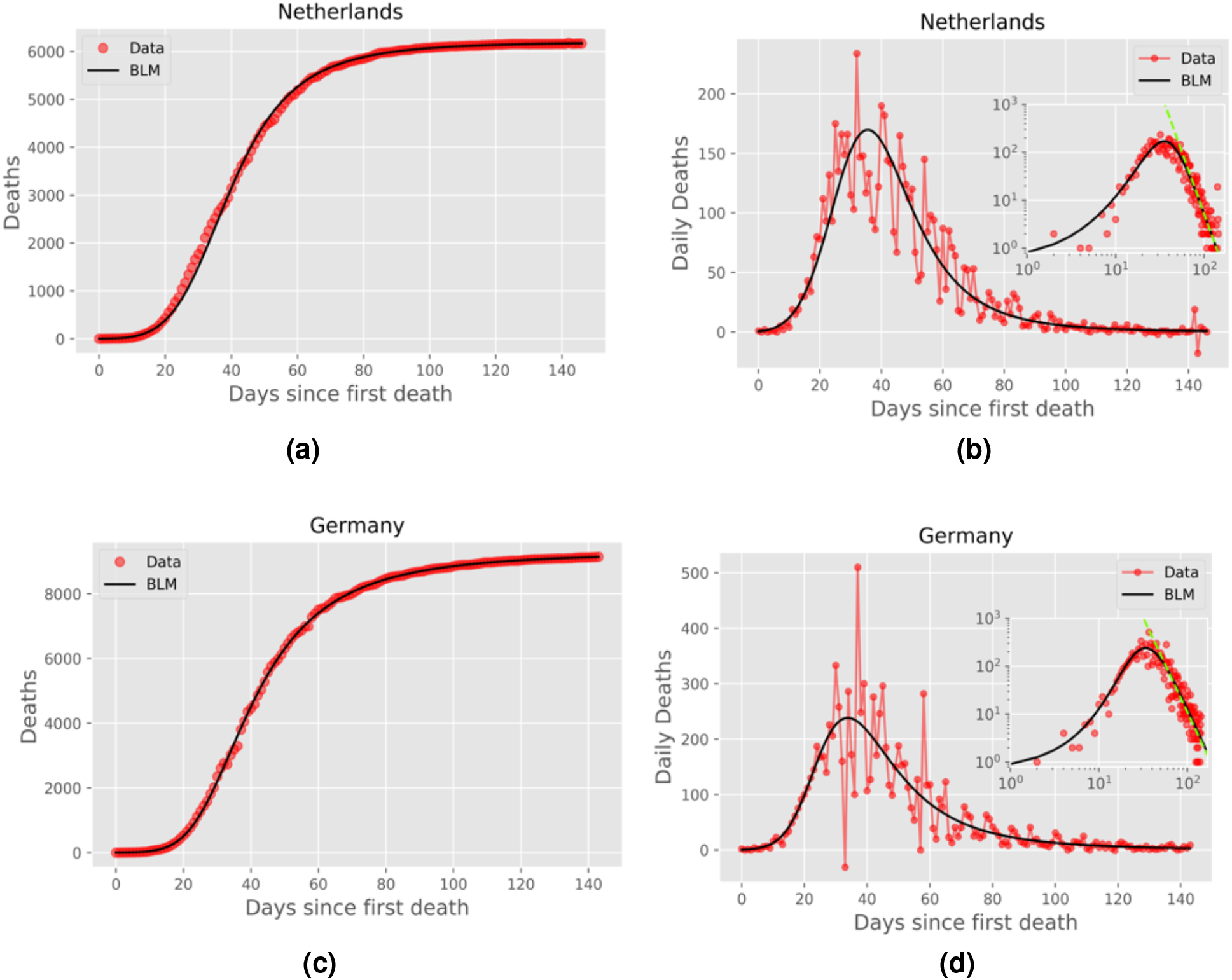
Same as in Fig. 2 for Netherlands and Germany.

In the right panels of Figs. 2-5 we show the daily numbers of deaths for the selected countries, where the red circles represent the empirical data and the black solid curves correspond to the time derivative of the theoretical curve *C*(*t*) predicted by the BLM, as obtained from the fits shown in the left panels of the respective figures. The insets in these figures show the same data as in the respective main plots but in log-log scale. Recall that a polynomial slow approach to the plateau for the cumulative number *C*(*t*) implies that the growth rate (i.e., the daily curve) *Ċ*(*t*) decays as a power law for large *t*. To highlight this power-law regime, we also show in the insets of the right panels of Figs. 2-5 the theoretical asymptotic power law (green straight lines), as predicted by Eq. (5). (We recall that a power law decay appears as a downward straight line in a log-log scale.)

Recall that the closer the exponent *p* is to 1, the closer the long-time asymptotic regime of the daily curve is to an exponential decay, and so the steeper the corresponding straight line in the insets of Figs. 2-4. Conversely, larger values of *p* indicate a slow power-law decay of the daily curve and hence a correspondingly less steep straight line in log-log scale. In other words, as *p* increases from nearly 1 to larger values, the power law (in log-log scale) goes from a nearly vertical straight line to a increasingly more horizontal one. This explains the behavior seen in Figs. 2-5, where in cases of countries with *p* rather close to 1, such as Canada and Italy (see Fig.2), the empirical daily curve approaches the power-law asymptotic behavior from the left; whereas for countries with larger *p*, such as Belgium and France (see Fig.4, the empirical curve approaches the power law from the right. Notably, for intermediate values of *p* (in the range 1.24-1.34), the daily curve reaches a near power-law behavior soon after the peak, as in the cases of Netherlands and Germany (see Fig.3).

Notice, furthermore, that even in situations where *p* is too close to or significantly higher than 1, the empirical data still tend to follow the power-law trend, as seen in Figs. 2 and 4. In other words, the effect of a power law regime predicted by the BLM is clearly noticeable in the real data for all cases analyzed here, even for cases where the data points do not lie on top of the asymptotic regime. We shall argue later (see Discussion) that the value of the exponent *p* might be seen as an indication of the effectiveness of the interventions adopted by a particular country to bring the epidemic under control. In this sense, a lower (higher) value of *p* is a reflection of more (less) effective measures, which thus leads to a faster (slower) approach to the saturation plateau for the cumulative curve.

**Figure 4.**
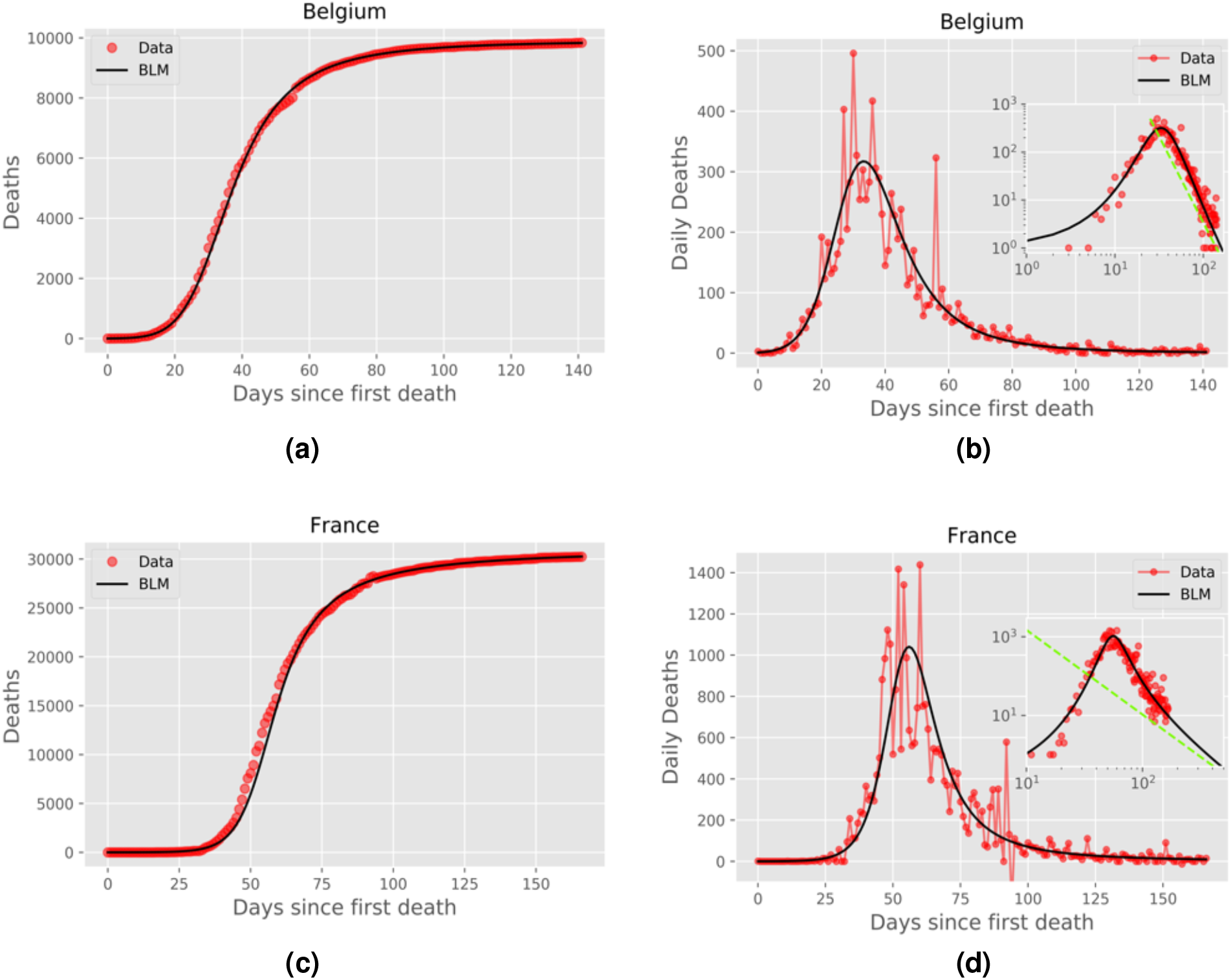
Same as in Fig. 2 for Belgium and France.

In Fig. 5 we show the cumulative and daily death curves for the Unites States up to July 7, 2020. One can see from this figure that the cumulative curve was trending toward a possible plateau, but it had not reached the same level of saturation as the other countries in Figs. 2-4. This slower approach to the plateau for the US is confirmed by the fact that it has the largest value of the exponent *p* among the countries considered here, as seen in table 1. Furthermore, the fact that power-law saturation regime was not quite well established in the US is also reflected in the large 95% CI for the exponent *p*. In other words, being further away from the plateau makes it more difficult to estimate the exponent *p* with good precision. As more data are accumulated, a better estimate for *p* should be obtained. Unfortunately, however, shortly after the date used for Fig. 5, the cumulative curve for the US underwent a change of trend, with the curve accelerating again, thus rendering the BLM unsuitable for subsequent times; see below for further discussion on this topic.

**Figure 5.**
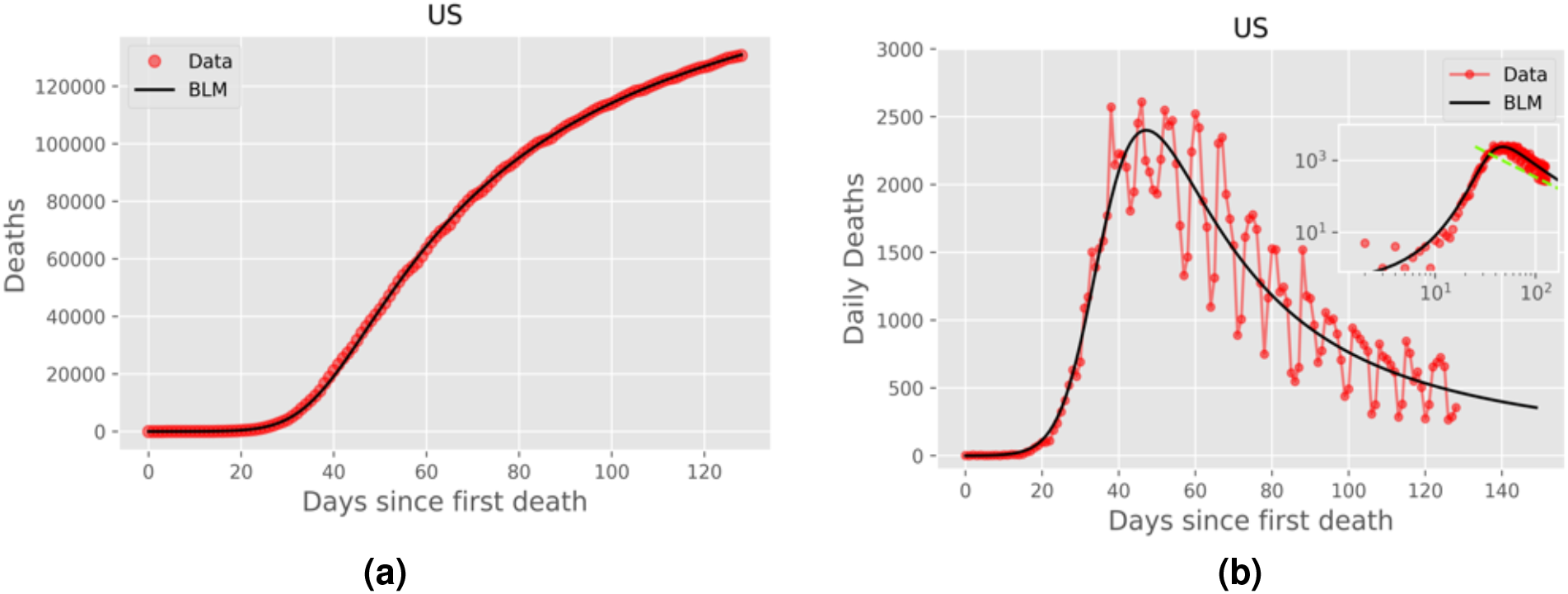
Same as in Fig. 2 for the United States, but here the data is up to July, 07, 2020.

We have seen above that the BLM is a very flexible model which is capable of describing epidemic curves with quite distinct profiles: in all cases shown in Figs. 2-5 the model predicts very well the entire dynamical evolution of the fatality curves within the period considered. Indeed, we have checked that the BLM, where *p* is a free parameter, yields better fits to all epidemic curves presented in Figs. 2-5 than the next less complex model, namely, the generalized Richards model (GRM) which fixes *p* = 1. In other words, the standard figures of merit implemented in the *lmfit* package for Python^22^ indicated better fits for the BLM in comparison to the GRM. This is further confirmation of the fact that the power law trend (i.e., *p >* 1) seen in the empirical data is a real dynamical feature.

In cases where the epidemic curve is either in the early growth phase or has not clearly entered a saturation regime, the BLM is of course not appropriate, so that an attempt to fit the empirical data with formula (2) will either not converge (former case) or produce unreliable estimate for the tail exponent *p* (latter situation). In such cases, one can use other simpler growth models, such as the generalised growth model (*p* = 0) the Richards model (*q* = *p* = 1), or the generalized Richards model (*p* = 1), to gain some insight into the early to intermediate stages of the disease^6^. Once the epidemic reaches a later phase, the BLM should then be used for a more complete description of the full evolution. (An automated version of this approach is currently available via an online app^23^.) In situations, however, where a ‘second wave’ of infection develops, no simple model, be it a growth model or a compartmental model, can describe the entire epidemic curve. Such more complex curves require, for instance, models with time-dependent parameters that can effectively capture both the first and the second phases of the dynamics^24^. Further discussion of second wave effects is beyond the scope of the present paper and will be left for a future publication^24^.

## 3 Discussion

We have seen above that the cumulative death curves of COVID-19 for many countries exhibit a slow, subexponential approach to the plateau, contrarily to the exponentially fast saturation predicted by most epidemiological models, such as mechanistic models of the SIR type. It is thus reasonable to argue that this slower than exponential saturation of the COVID-19 dynamics stems from the complex human response in the late phase of the epidemics. It is normally the case that countries impose stringent containment measures in the early and intermediate stages of the epidemics, which are then progressively relaxed once the worst phase of contagion has been left behind. It is therefore expected that countries that have kept stricter measures, of one sort or another, for longer periods should experience a faster approach to the end (plateau) of the epidemics; whereas countries with more relaxed sets of measures should display the opposite trend (slow saturation regime).

This assumption is largely supported by the data analysis reported above. For instance, Italy, which enforced one of the longest and most stringent lockdown, has one of lowest values of *p*, indicating a fast, near-exponential saturation onto the plateau. Canada, which has a value of *p* comparable to Italy’s, also instituted strong lockdown measures in March, 2020. Other European countries, besides Italy, also adopted lockdown measures, but they “closed less and reopened earlier”^25^, which might help to explain why, say, France has a considerably larger *p* than Italy’s. The Netherlands and Germany have a somewhat intermediate *p* value—significantly larger than Italy’s but smaller than that of France’s. These two countries did not adopt a conventional lockdown, but implemented a consistent set of measures to enforce social distancing (among other control measures)^26,27^, which may have lead to a reasonably small *p*. It is important to emphasize, however, that the exponent *p* characterizes only the final part of the epidemic curve and hence it is not strongly related to how well countries have been able to control the epidemic in terms, say, of mortality rates. In other words, countries that have low mortality rates can nonetheless display comparatively higher values of *p*; and vice-versa.

The total number of fatalities that occur in the late phase of the epidemic is, of course, quite smaller than that in the early and intermediate stages. Nonetheless, it is important for authorities to aim at a high ‘rate of approach’ to the plateau (i.e., small *p*), because the longer the virus lingers on in the population, the greater the risk of a resurgence of infections, which may in turn prolong even more the epidemic. More to the point, once countries have managed to ‘bend the curve’, they should try to climb to the plateau as close to exponentially fast as possible. Failing to do that may significantly increase the total number of victims at the end of the pandemic. It is also worth noting that the subexponential growth recently documented in COVID-19 epidemic data has been usually attributed to the early adoption of mitigation measures^6, 28, 29^. On the flip side of the coin, the subexponentially slow approach to the plateau that we have reported here seems to be, in part, a consequence of relaxing these measures. It is also reasonable to argue that the slower the approach to the plateau, the greater the risk of a subsequent resurgence of infections. Indeed, at the time of the revision of this paper, most of the countries discussed here have shown signs of a second wave of infections, whereby their epidemic curves have re-accelerated. Such more complex dynamics is of course beyond the capabilities of the growth models considered here, which display only one inflection point. One possible generalization of the present work, aimed at capturing second wave effects, is to allow the parameters of the BLM to vary in time. We are currently investigating this interesting possibility.

## 4 Conclusion

The relevance of epidemiological models often lies in the dynamical regimes and scenarios they are able to predict and/or explain, rather than in the specific values of the parameters they produce. In this perspective, the main contribution of the present paper is to predict a novel behaviour for the COVID-19 epidemic, namely: a polynomially slow approach of the growth curve toward the plateau at the end of the epidemic. This implies in turn that the rate of growth (i.e., the daily number of deaths) decays in time, after the peak, as a power law, contrarily to the exponentially fast decay predicted by standard compartmental models.

Here we have modelled the fatality curves of COVID-19 by means of a generalised logistic model that is able to capture not only a possible subexponential early growth regime but also a subexponential saturation dynamics in the final phase of the epidemics. In particular, our model shows that the ‘residual’ number of deaths, defined as the difference between the expected final death toll and the current number of deaths, decays in time as a power-law. This implies, equivalently, that the daily number of deaths exhibits a power law decay in the final phase of the epidemic (if second-wave effects do not come into play). We have furthermore argued that the less strict the control measures in the late phase of the epidemic, the smaller the respective power-law exponent and hence the slower the approach to the plateau. This, in turn, implies a greater risk of a resurgence of infections, as the virus continues to propagate in the population (even if at a slower pace) for a longer period of time. Although here we have restricted ourselves to mortality data, it is expected that a similar power-law behaviour should also hold for epidemic curves of confirmed cases.

The results reported in the present paper are relevant both from a basic viewpoint, in that they show that additional care is needed when modelling the terminal phase of epidemics, and from a practical perspective, for they remind policymakers and health authorities that it is important not only ‘to bend the curve,’ but also to make sure that it reaches its dynamical end as quickly as possible.

## Methods

### Mathematical Model

As indicated before, in this paper we model the fatality curves of the COVID-19 epidemic via the growth model defined in Eq. (1). This equation must be supplemented with an initial condition, which we take as follows:

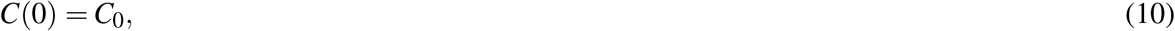

for some given value of *C*_0_. The model defined in (1) is one of the most general growth models in that it recovers, as particular cases, several other known models, namely: i) the logistic model (*q* = *p* = *α* = 1); ii) the Richards model (*q* = *p* = 1); iii) the generalized Richards model (*p* = 1); iv) Blumberg’s hyperlogistic equation (*α* = 1); v) the generalised logistic model (*p* = *α* = 1); and vi) the generalised growth model (*p* = 0).

Equation (1) was introduced by Tsoularis and Wallace^16^ and was referred to by these authors as the generalised logistic function. However, the term generalised logistic model has been more commonly used in connection with the particular case *α* = *p* = 1^30^. Here we propose instead the terminology *beta logistic model* (BLM), because the right-hand-side of (1) corresponds precisely to the generalised beta distribution of the first kind^31, 32^, which is defined by the following probability density function (pdf):

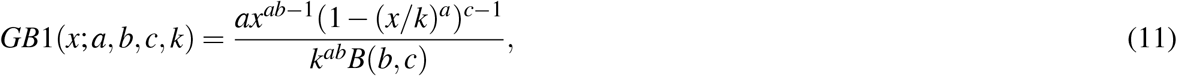

for 0 < *x* < *k* and *a*;*b*;*c*;*k* > 0, where *B*(*b*;*c*) is the beta function defined as

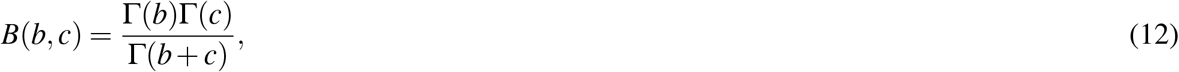

with Γ (·) denoting the gamma function. In view of (11), Eq. (1) can be rewritten as

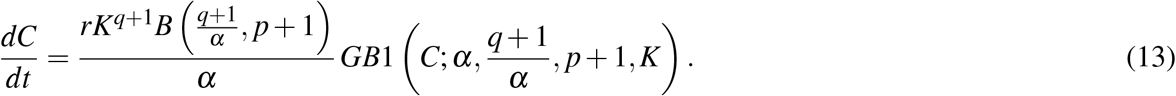

Viewing the right-hand side of (1) as a pdf is useful to gain some insights into the underlying growth dynamics. Seen in this perspective, the model (1) encodes the fact that the rate of growth of the disease (be it measured in number of cases or deaths) at a given time *t* is proportional to the product of two terms: the first term represents the current ‘strength’ of disease, thus being related to the number of cases in the population; and ii) the second term accounts for the available ‘space’ in the population for the disease to grow. Note, however, that in contrast to the standard logistic model where both such product terms are linear in *C*(*t*), i.e., *q* = *p* = *α* = 1, the BLM seeks to take into account, albeit in an effective and indirect manner, the fact that the virus propagates in a complex network of human contacts, which in turn is affected by the very measures adopted to curtail the spread of the disease. This complex dynamics is captured in the BLM via the introduction of the exponents *q, p*, and *α*, which results in the generalised beta distribution.

It turns out that the model (1) can be integrated exactly. To see this, we start by introducing the reduced number of cases

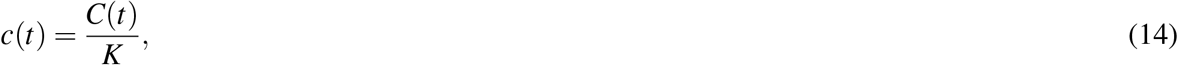

so that (1) becomes

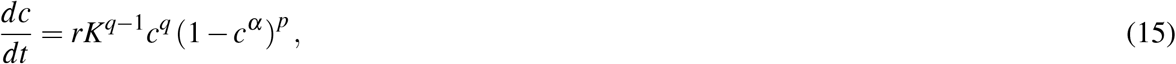

After a trivial separation of variable, we write

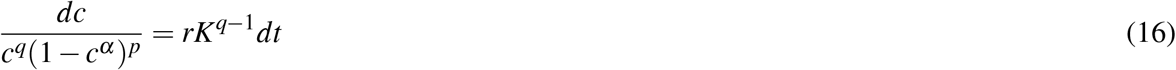

which upon integration on both sides yields

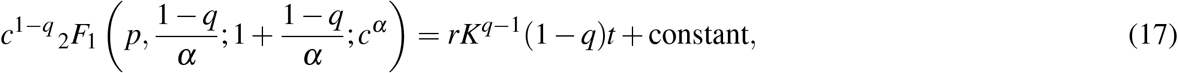

where _2_*F*_1_(*a, b*; *c*; *x*) is the Gauss hypergeometric function^20^. After fixing the constant of integration with (10), we obtain

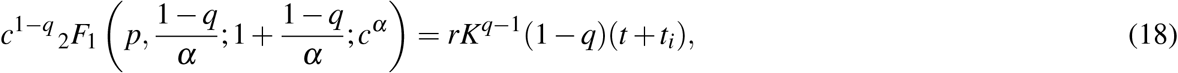

Where

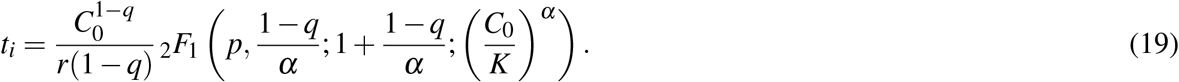

Combining (18) and (19), and returning to the original variable *C*(*t*), we obtain the solution given in (2).

The BLM as defined in (18) has two ‘extensive’ parameters, namely *C*_0_ and *K*, which represent the initial and final number of cases, respectively, and four ‘intensive’ parameters, namely: *r, q, α*, and *p*, which are related to the intrinsic dynamics of the epidemic in each particular group of individuals. It is therefore instructive to seek a better understanding of the dynamical (and epidemiological) roles that these parameters play.

The growth rate *r* sets the time scale of the problem, and as such it reflects the epidemiological conditions in the early growth phase of the epidemic. In other words, *r* is expected to be directly related to the basic reproduction number, *R*_0_, since the higher the *R*_0_, the faster the virus spreads and the larger the growth rate *r* should be. (Broadly speaking, *R*_0_ can be defined as the expected number of secondary infections caused by a primary infectious individual in an entirely susceptible population.)

The parameter *q*, as already anticipated, is related to the nature of the growth regime (if exponential or subexponential) in the beginning of the epidemic. To see this, first note that if *C*≪*K*, the ODE (1) yields the so-called generalised growth model^30^:

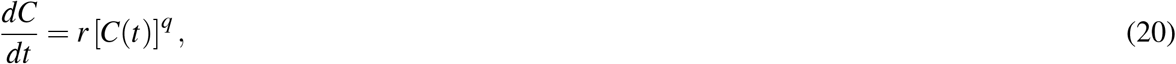

whose solution is simply

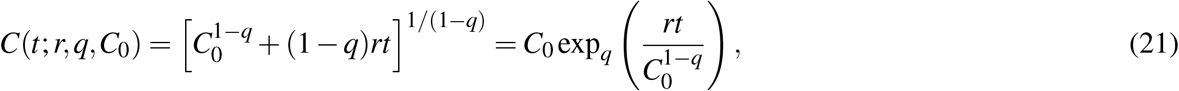

where the function exp_*q*_ (*x*) = [1 + (1 *−q*)*x*]^1*/*(1*−q*)^ is known in the physics literature as the *q*-exponential function^33^. The parameter *q* thus characterises the early growth dynamics, allowing the model to include from a linear growth (*q* = 0) to a subexponential regime (0 *< q <* 1) and to a fully exponential growth (*q* = 1). (Values of *q* greater than one in (21) are ruled out, because they would lead to a superexponential spread of the disease, which is neither epidemiological reasonable nor mathematically acceptable, as it would lead to a singularity in finite time.) In particular, note that according to (21) the early growth of the epidemic curve is approximately polynomial for 0*≤q <* 1, as given in the top equation in (4). Polynomial early growth has been observed across a range of epidemics before^13^. It has also been recently identified in the COVID-19 data for several countries^6, 28, 30^.

In the intermediate region of the epidemic curve, one observes an approximately linear regime around the inflection point *t*_*c*_, with a small third-order deviation from a straight line. More specifically, from (1) one readily finds that the expansion of *C*(*t*) about *t* = *t*_*c*_ is given by

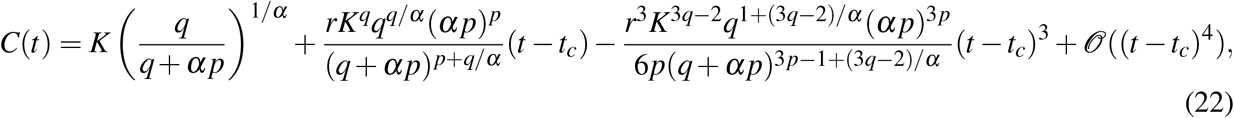

which corresponds to the expression given in (6)–(9).

Finally, in the long-time asymptotic limit, i.e., when *C*(*t*) *→ K*, we can investigate the behaviour of the BLM curve by introducing the small quantity *ε*(*t*) = *K −C*(*t*). Inserting this definition in (1) and considering *ε* ≪ 1, we obtain to leading order:

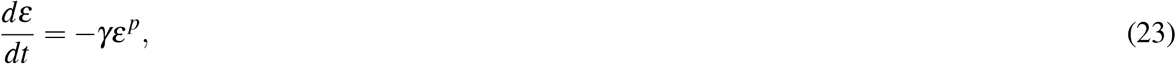

where *γ* = *rα*^*p*^*/K*^*p−q*^, which upon integration yields

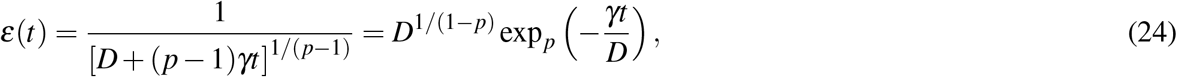

with *D* denoting an arbitrary constant of integration. This shows that in general the BLM curve *C*(*t*) approaches the plateau *K* according to a *p*-exponential function (i.e., the *q*-exponential function defined before with *q* = *p*). [Eq. (23) also appears, e.g., in the problem of protein folding/unfolding, leading to *q*-exponential relaxation phenomena there as well.^34^]. In particular notice that, for sufficiently large times *t*, the curve *C*(*t*) generically approaches the plateau polynomially slowly in time, in the sense that *ε*(*t*) decays in time as a power law:

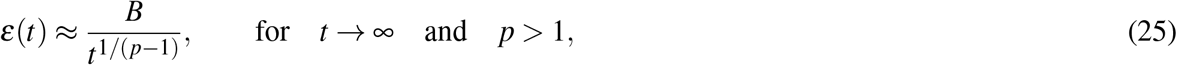

*B* = [*K*^*p−q*^*/*(*p−* 1)*rα*^*p*^]1*/*(*p−*1), which recovers the bottom equation in (4). In contrast, only for *p* = 1 does one get an exponentially fast saturation into the plateau: *ε*(*t*) ∝ exp(*−γt*). It follows from (25) that the growth rate asymptotically decays as a power-law for large times:

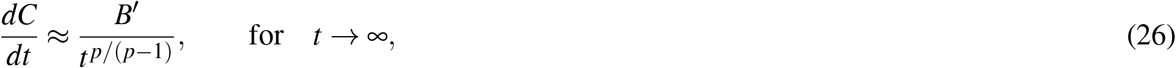

where *B′* = *B/*(*p−* 1). Eq. (26) thus shows that the corresponding daily curves also display power-law decay for large times. In this context, it is worth recalling that compartmental models of the SIRD type predict an exponential decay for the rates of both new infections and deaths toward the end of the epidemic.

### Data Availability and Statistical Fits

Here we first briefly note that because the number of confirmed cases is a poor metric for the advance of the COVID-19 epidemic, owing to the fact that a large proportion of infections go undetected, we have adopted the fatality curves as our primary objects of study, since it is widely recognised that the death count is a more reliable statistics in this case^6^. The data used in this study were obtained from the database made publicly available by the Johns Hopkins University^35^. We have used data for the total number of deaths up to July 31, 2020, for the following countries: Canada, Belgium, France, Germany, Italy, Netherlands, and the United Kingdom; whereas for the United States we used data up to July 07, 2020.

To perform the statistical fits reported in this paper, we employed the Levenberg-Marquardt algorithm to solve the non-linear least square optimisation problem, as implemented in the *lmfit* package for Python, which not only provides parameter estimates but also determines their respective confidence levels. Technically, because the exact solution for the BLM is given in implicit form, we have to fit the analytical formula given in (2) to the empirical datasets seen as curves of the type *t*(*C*). But this imposes no difficulty in the numerical routine whatsoever. To reduce the number of free parameters in the model we have set *C*(0) = *C*_0_, where *C*_0_ is the number of deaths recorded at the first day that a death was reported, so that we were left with five parameters, namely (*r, q, α, p, K*), to be determined numerically. Growth models with several parameters are known to be susceptible to the problem of overfitting and parameter redundancy^6, 17, 30, 36^. To minimise this risk we have adopted some precautions, as discussed next.

First, we limited the exponent *q* to the mathematically and epidemiological acceptable range 0 *≤ q≤*1. When the *lmfit* routine returns *q* = 1, it usually cannot estimate the confidence levels because this is the upper bound of *q*; in this case we assume that *q* = 1 is an ‘exact value’ for the country in question. As for the remaining parameters, no restriction was enforced *a priori* other than their natural ranges (*r, α, K >* 0 and *p >* 1). Nevertheless, we checked the estimates returned by the curve-fitting routine against possible inconsistencies. For example, the growth rate *r* is expected to be typically within the interval (0,1)^6^ and that has been the case for all countries considered here. Similarly, the asymmetry parameter *α* is typically smaller than 1^6^, but countries that display a somewhat ‘sharp’ bending after the inflexion point (such as France, see Table 1) are expected to have higher values of *α*. In the same spirit, we checked the values of the exponent *p*, which determines the power-law decay of the growth rate *Ċ*(*t*) given in (5), by comparing the tails of *Ċ*(*t*) for both the empirical data and the theoretical prediction. Good agreements between the BLM and the data, over the entire, including the tails, were observed in all cases reported here, which validates the estimates of the model parameters.

As already indicated, the computer codes for the statistical fits were written in the *Python* language, and the plots were produced with the data visualisation library *Matplotlib*. All numerical codes and corresponding data sets used in this paper are available in our group’s software repository at fisica. ufpr.br/redecovid19/software or can be requested from the authors. The fitting-curve routine for the BLM can be automatically tested for other countries (not included here) via an app available online^23^.

## Data Availability

The dataset is compiled from data provided by Johns Hopkins University at https://data.humdata.org/dataset/novel-coronavirus-2019-ncov-cases.

https://fisica.ufpr.br/modinterv

